# Detection of COVID-19 and age-dependent dysosmia with paired crushable odorant ampules

**DOI:** 10.1101/2022.03.13.22271253

**Authors:** Ronald W. Wood, Christopher J Stodgell, Mitchell A. Linder, Eva K. Pressman

**Affiliations:** Department of Obstetrics and Gynecology, University of Rochester School of Medicine and Dentistry, Rochester, New York, United States of America; Department of Urology, University of Rochester School of Medicine and Dentistry, Rochester, New York, United States of America; Department of Neuroscience, University of Rochester School of Medicine and Dentistry, Rochester, New York, United States of America

## Abstract

**Background:** Signs of anosmia can help detect COVID-19 infection when testing for viral positivity is not available. Inexpensive mass-produced disposable olfactory sensitivity tests suitable for worldwide use might serve not only as a screening tool for potential infection but also to identify cases at elevated risk of severe disease as anosmic COVID-19 patients have a better prognosis.

**Methods and Findings:** We adopted paired crushable ampules with two concentrations of a standard test odorant (n-butanol) as standard of care in several clinics as community prevalence of COVID-19 infection waxed and waned. This was not a clinical trial; a chart review was undertaken to evaluate the operating characteristics and potential utility of the test device as RT-PCR testing became routine. The risk of anosmia was greater in COVID-19 patients. Olfactory sensitivity was concentration-dependent, decreased with aging, and was sex-dependent at the highest concentration. Hyposmia was detected across a wider age range than expected from the literature, and tests can be optimized to characterize different age groups.

**Conclusions:** n-Butanol at 0.32 and 3.2% in crushable ampules can be used to characterize olfactory function quickly and inexpensively and thus has potential benefits in pandemic screening, epidemiology, and clinical decision-making.

## Introduction

Olfactory dysfunction is a well-documented early sign of SARS-CoV-2 infection. Advancing age is correlated with olfactory dysfunction (1); but older COVID-19 patients have a lower prevalence of olfactory dysfunction (2), and the best predictors of death in hospitalized patients are age and a lack of anosmia (3). Conversely, COVID-19 patients with dysosmia have less severe symptoms (4, 5), are younger (6, 7), and have a better prognosis (3, 8-15); however, this may not hold for cases with prolonged anosmia (16-18). Obstruction of the olfactory cleft by swollen tissues of the olfactory mucosa results in anosmia (19), thus sudden, unexplained onset of anosmia likely is an indicator of the intensity of the initial inflammatory immune response to viral infection.

Olfactory dysfunction is detected more frequently when objective measurements are used instead of self-reports (2, 9, 20-22). Odor identification tests require familiarity with odors, are culturally dependent (23) and require language competency (24-29). Unlike “scratch and sniff” type tests for olfactory dysfunction that release suprathreshold stimuli and require identification of the odorant among several choices (30), the use of olfactory stimuli at graded concentrations assess sensitivity directly (9, 31). However, commercially available “gold standard” systems tend to be relatively expensive, time consuming, and pose risks in a pandemic as they are intended to be reused and may act as fomites for disease transmission (32). Disposable odorant detection tests that are instantaneous, requiring a “yes” or “no” response seconds after presentation, and, in pandemic times when global testing is needed, should have minimal costs so that large scale use is feasible.

## Methods

We undertook a retrospective chart review of patients who received routine objective measurement of olfactory function by determining if they could detect two concentrations of n-butanol (initially 0.06 and 3.2% in water) packaged in conjoined crushable ampules containing 0.3 ml of the test odorant (S1 Fig). The higher concentration of n-butanol (3.2%) was selected because it is suprathreshold for normal individuals (33); the initial low concentration (0.06%) was selected in an attempt to identify the 12.4 percent of a normal population over the age of 40 expected to be hyposmic (1). Test concentrations remained the same at Preoperative Site 1 as were used in the Respiratory Clinic, however, the low-concentration ampule was increased to 0.32% n-butanol at Preoperative Site 2. One tip of the test unit was marked yellow to denote the low concentration; yellow or blue food color was added to the odorant and thus changed the appearance of each ampule when crushed. The two ampules were joined with polyolefin shrink tubing. The URSmellTest ™ was incorporated as an intake procedure at three clinics after the onset of the COVID-19 pandemic. A laminated instruction sheet was used by clinic staff to standardize the testing procedure (S2 Fig).

The UR Primary Care Central Respiratory Clinic was established to direct patients at risk of COVID-19 (i.e., individuals who were presenting with clinical symptoms of SARS-CoV-2 infection) to a site where high level precautions were implemented at a time when personal protective equipment availability was limited. Results were recorded in the electronic medical record system from 14 Dec 2020 to 7 Jan 2021 during which 225 olfactory tests had accompanying RT-PCR results. The mean percent viral positivity in Monroe County New York during this interval was 9.2 ± 1.35 percent (mean ± standard deviation), but was 31 percent at this clinic, roughly 3 times higher than the county prevalence. After closing of the UR Primary Care Central Respiratory Clinic once the late 2020-early 2021 surge subsided and elective procedures resumed, use of this test continued at two successive preprocedural test sites (*Preoperative Site1* and then subsequently *Preoperative Site 2*) that performed SARS-CoV-2 testing on all patients presenting for procedures requiring admission to Strong Memorial Hospital, resulting in better representation of the community in terms of age distribution (Table 4) and demographics (S1 Table) than the patients presenting with respiratory symptoms at the height of the midwinter surge. Viral prevalence at these sites was lower in this clinical context compared to the community prevalence, presumably because individuals known to have active COVID-19 infection were instructed to stay home and reschedule their elective procedures.

Chart review protocol was approved by the University of Rochester Research Subjects Review Board: Study00006033: Dysosmia Chart Review.

## Results

### Respiratory clinic site

There were no sex differences observed for age or viral positivity (χ^2^ _(1df)_ = 1.95, p=0.163). Viral infection was associated with highly significant effects when the frequency of anosmia was compared to those with normal function (χ^2^_(1df)_ = 9.97, p=0.0016).

Odds ratios provide a measure of the strength of association between exposure and an outcome, representing the odds that an outcome will occur in the presence of SARS-CoV-2 exposure compared to those in the absence of viral infection (OR=3.36, p=0.002, two tailed; Table 1). Relative risk is the ratio of the probability of an outcome in the two groups. The risk of anosmia was 2.40 times greater if a patient displayed SARS-CoV-2 positivity (p= 0.0016, two tailed; Table 1).

**Table 1.** Odds and relative risk ratios of anosmia using n-butanol (3.2%) detection in a respiratory clinic setting during a COVID-19 surge.

| Odds ratio |  |  |  |  |  |
| --- | --- | --- | --- | --- | --- |
| <u>Odds</u><br><u>ratio</u> | <u>S</u><br><u>E</u> | <u>Z</u> | <u>95% confidence limits</u> |  | <u>p</u> |
| 3.36 | 0.<br>394 | 3.07 | 7.26 | 1.55 | 0.00212 |
| Relative risk ratio |  |  |  |  |  |
| 2.40 | 0.<br>279 | 3.15 | 4.15 | 1.39 | 0.00164 |

Comparator data on the frequency of dysosmia in the population comes from the National Health and Nutrition Examination Survey (NHANES) study. NHANES estimated the prevalence of olfactory dysfunction in the US population over the age of 40 (1), providing estimated proportions in ten-year age brackets. These were used to provide an estimated proportion likely to show olfactory dysfunction based on the age distribution in this sample. Patients from the UR Primary Care Central Respiratory Clinic were a symptomatic respiratory clinic population and not a normal healthy population sample like the NHANES data. The mean age of the patients seen in the clinic was 54.3 years, standard deviation 16.2. The median age of the respiratory clinic patients was 55 years; although olfactory dysfunction normally increases with age, in COVID-19 patients increasing age is correlated with lower prevalence of olfactory dysfunction (2), and lack of anosmia is associated with higher risk of severe illness and death in hospitalized patients(3, 14, 15). Uninfected patients in the sample of patients from this site and over the age of 40 had 1.65 higher than expected incidence of anosmia; SARS-CoV-2 infection was associated with anosmia incidence ∼3.9 times higher than expected from the NHANES study (Table 2). This simple ratio of risks is 2.39, which is consistent with the above relative risk estimate (2.40) without attempted age correction. Thus, despite the lower incidence of anosmia in severe COVID-19 disease with respiratory complications, these findings robustly demonstrate that crushable ampules can be used to detect COVID-19-associated anosmia in clinical settings.

**Table 2.**
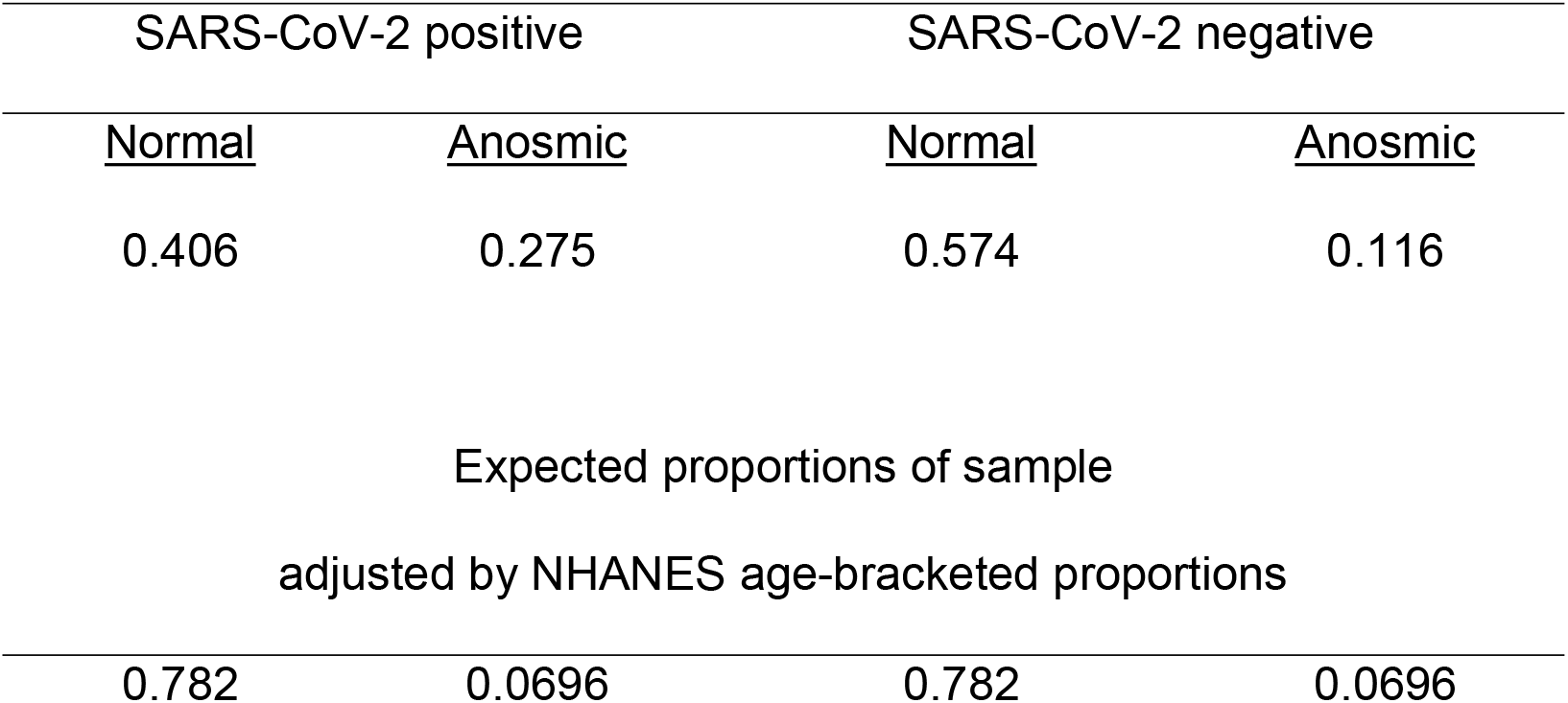
Age adjusted estimates of the proportions based on NHANES prevalence estimates. Observed proportions of patients presenting at the Respiratory Clinic

| SARS-CoV-2 positive |  | SARS-CoV-2 negative |  |
| --- | --- | --- | --- |
| <u>Normal</u> | <u>Anosmic</u> | <u>Normal</u> | <u>Anosmic</u> |
| 0.406 | 0.275 | 0.574 | 0.116 |
| 0.782 | 0.0696 | 0.782 | 0.0696 |

### Preoperative Site 1

Viral prevalence was very low in this context and in the period from Jan 29, 2021 to May 27, 2021. Although there was a relationship to age by logistic regression (Fig 1, Table 3) using Newton’s method (34, 35), the 0.06% n-butanol concentration was not easily detected even by staff and younger SARS-CoV-2 negative patients (n=905); thus, a higher concentration (0.32%) was used subsequently.

**Table 3.** Logistic regression model coefficients and constants.

|  | Hyposmia |  | Anosmia |  |
| --- | --- | --- | --- | --- |
|  | Site 1 (0.06%) | Site 2 (0.32%) | Site 1 (3.2%) | Site 2 (3.2%) |
| $\beta_0$ (SE) | -1.7377 (0.2588) | -2.4059 (0.2407) | -5.5046 (0.8614) | -5.3171 (0.5782) |
| $\rho$ | <0.0001 | <0.0001 | 0.0166 | <0.0001 |
| $\beta_1$ (SE) | 0.0133 (0.0044) | 0.0198 (0.0041) | 0.0330 (0.0133) | 0.0395 (0.0090) |
| $\rho$ | 0.0025 | <0.0001 | 0.0130 | <0.0001 |
| $X^2$ | 9.37 | 22.64 | 6.91 | 22.95 |
| $\rho$ | 0.0022 | <0.0001 | 0.0086 | <0.0001 |
| $\eta$ | 905 | 1290 | 905 | 1290 |

| n-Butanol Concentration | Variable | Coefficient | Standard Error | p-value | Odds Ratio | 95% Confidence Interval |
| --- | --- | --- | --- | --- | --- | --- |
| 0.32% | Age | 0.0180 | 0.0042 | <0.0001 | 1.0182 | (1.0098,1.0267) |
|  | Sex | -0.1337 | 0.1435 | 0.3516 | 0.8749 | (0.6603,1.1591) |
|  | Constant | -2.2697 | 0.2727 | <0.0001 |  |  |
| 3.2% | Age | 0.0361 | 0.0091 | 0.0001 | 1.0367 | (1.0183,1.0554) |
|  | Sex | -0.6673 | 0.2706 | 0.0136 | 0.5131 | (0.3019,0.8719) |
|  | Constant | -4.7572 | 0.6124 | <0.0001 |  |  |

**Table 4.**
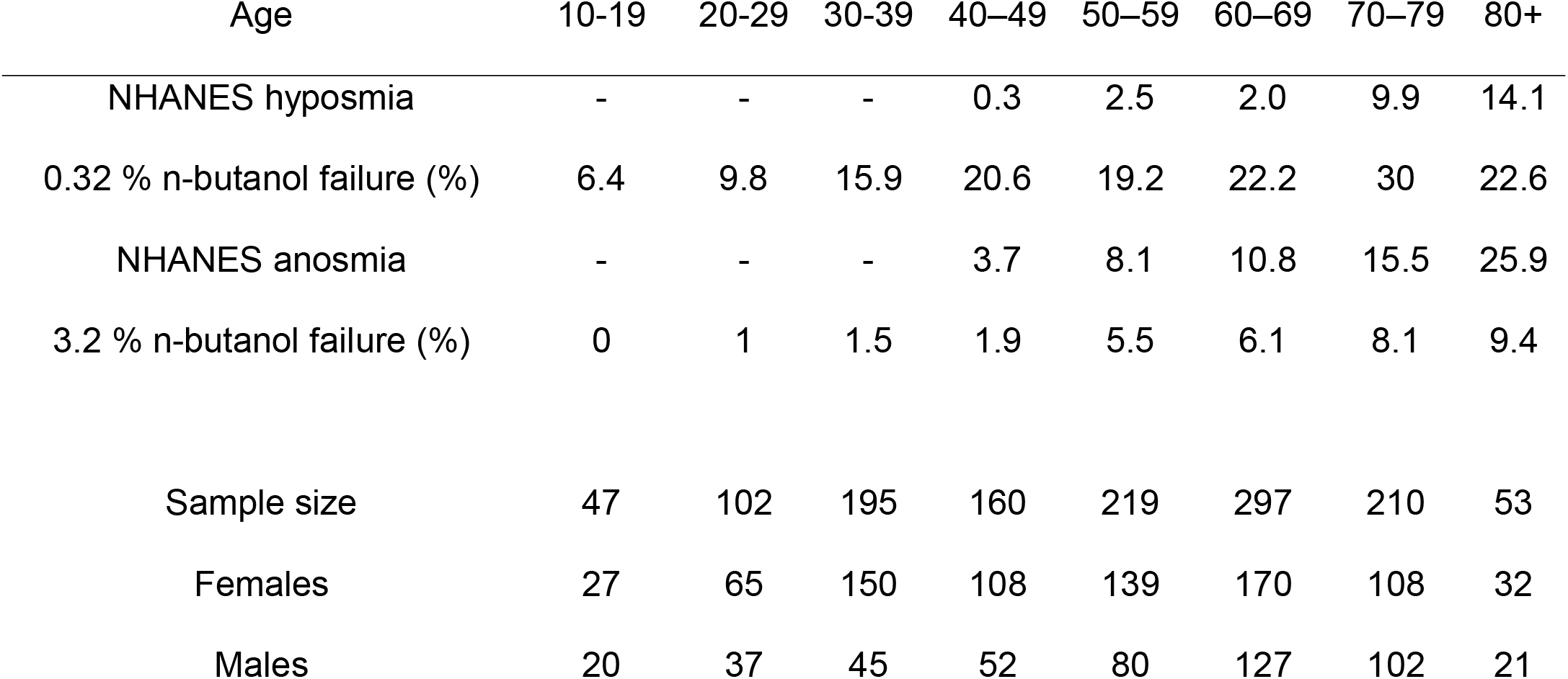
Age decade bracketed prevalence of hyposmia and anosmia at the second preoperative testing site for comparison with the NHANES findings(1) that relied on suprathreshold odorant identification error rates in SARS-CoV-2 negative patients. The present sample was drawn from the 1,308 preoperative clinic presentations, which were screened at the two sites, because there were 5 subsequent deaths in the interval of observation, 13 SARS-CoV-2 positive patients, and 7 patients less than age 10; the final distribution is based on the remaining 1,290 living SARS-CoV-2 negative patients.

**Fig 1.**
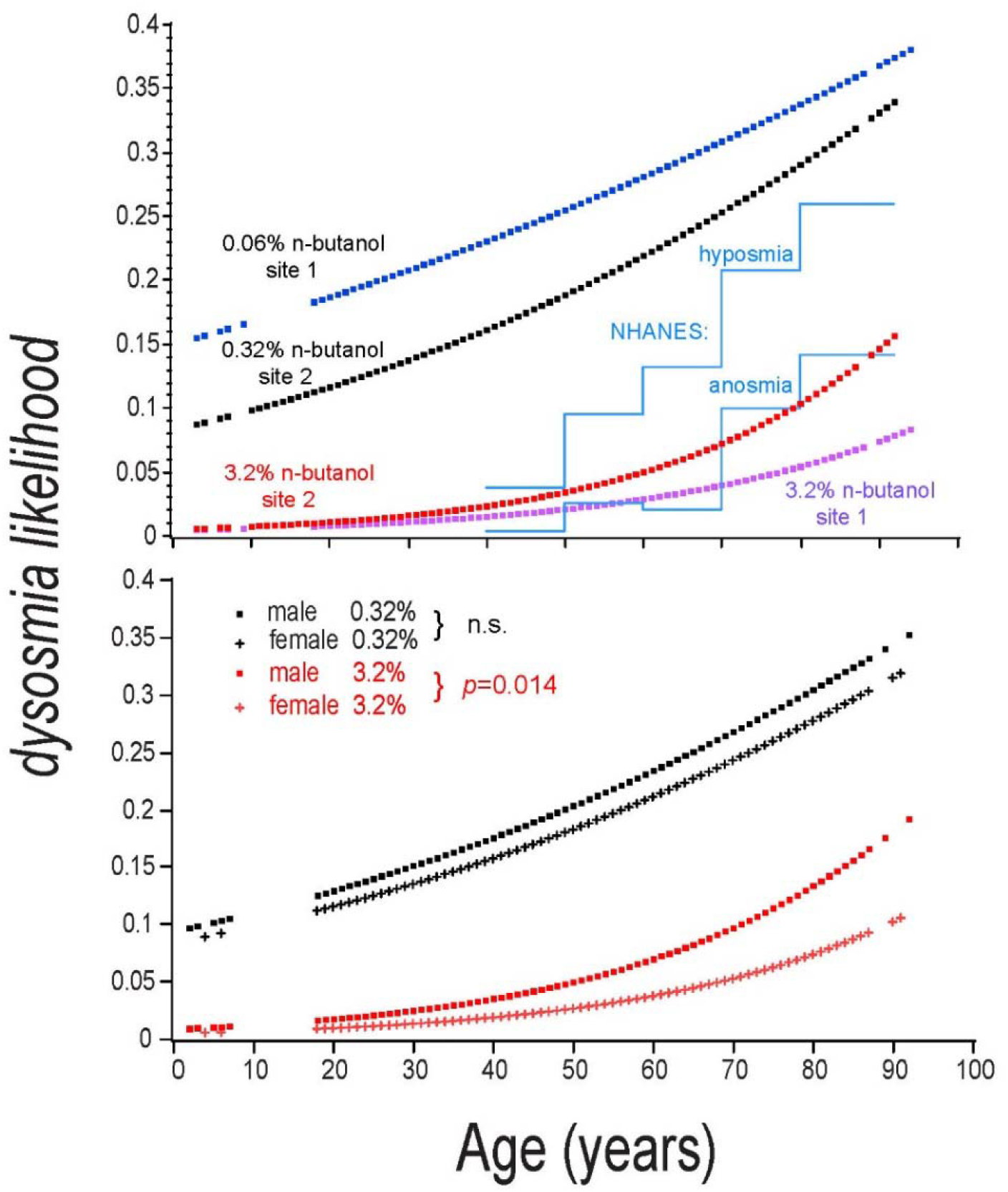
Logistic regression estimates of olfactory dysfunction as a function of age and concentration. Top panel: The likelihood of detecting the high concentration odorant was greater at Preoperative Site 1, where the first test stimulus was more difficult to detect (i.e., n-butanol concentrations of 0.06% vs 3.2%) Blue: 0.06% n-butanol in water at preoperative site 1; Black: 0.32% n-butanol at site 2; Red: 3.2% n-butanol at site 2; Purple: 3.2 % at site 1; Blue lines: NHANES(1) estimates based on odor identification error rates. Bottom panel: As females age they retain olfactory sensitivity to a greater extent than males at 3.2%, but not at 0.32%, thus there is a three-way interaction between age, n-butanol concentration, and sex on n-butanol sensitivity.

### Preoperative Site 2

Site 2 opened June 1, 2021 after Preoperative Site 1 closed, a simple relocation within the community operated by the same staff to manage the same queue of patients that were being seen at Preoperative Site 1 for scheduled procedures. All test materials from Site 1 were recovered and replaced with tests with the revised “lower” test concentration of 0.32% n-butanol in water. At this site 1,290 patients were tested until October 14, 2021. Logistic regressions as a function of age were significant (Table 3 top); point estimates are presented for comparison with the NHANES estimated prevalence based on odor identification errors (Fig 1 top panel). Logistic regression was performed with the addition of sex as an independent variable (Fig 1, lower panel; Table 3 bottom).

### Age and Sex determine Dysosmia Incidence

Age-dependent increase in n-butanol olfactory threshold has been reported (36) using time- and labor-intensive threshold determinations in individual subjects; we systematically replicated this phenomenon using a quick test that demonstrated a concentration-dependent loss of sensitivity across a wide age span (Fig 1). The NHANES study (1) utilized an eight-item scratch and sniff suprathreshold odor identification test (Sensonics International, Haddon Heights, NJ) that defined hyposmia and anosmia as bracketed error rates, and demonstrated increasing misidentification with age (Table 4 (1); solid blue lines in Fig 1). The two low n-butanol concentrations were more sensitive to olfactory impairment than the NHANEs criteria, but the high concentration odorant test was in reasonable agreement with the NHANEs data (Table 4; Fig 1 top).

Sex differences in n-butanol olfactory sensitivity have been reported (33, 37), but did not observe or report an interaction with age. We report a concentration-dependent interaction with sex, females retaining more sensitivity with age only when tested with the high concentration odorant, 3.2% n-butanol (Fig 1, lower panel).

## Discussion

The NHANES study (1) used odorant misidentification errors to identify hyposmia and anosmia; note that odorant misidentification errors do not necessarily mean the subject could not detect the odorant, as parosmia is a misidentification error, a known consequence of COVID-19 disease. This landmark study estimated the prevalence of measured olfactory dysfunction to be 12.4 % in adults 40 or more years of age. In the context of olfactory sensitivity thresholds, this would correspond to the tail of the threshold distribution approximately one standard deviation above the mean but is age and sex dependent. The lower of two test concentrations thus might be adjusted to identify the hyposmic across age ranges encountered in different educational or geriatric settings. Logistic regression on additional series of observations of test concentrations could be collected to establish nomograms, and subsequently used to in conjunction with census and other datasets to estimate the true burden of olfactory dysfunction. In the absence of such experimental work at a time of need, the present chart review provided an opportunity to provide interim estimates based on the logistic regression functions obtained at three concentrations and are reported in S2 Table. Note that this report demonstrates that the 0.32% concentration is sensitive to age-dependent changes in sensitivity, and in the context of the progression of COVID-19 olfactory dysfunction, impaired detection of low concentrations would occur before the onset of complete loss of smell and recover after the ability to detect higher concentrations returns.

Since this data arose from a chart review of a series of simple clinical tests of olfactory sensitivity, no experimental design considerations were in effect to control for any sequence or expectancy effects that we encountered after the fact. Patients were told that their sense of smell was going to be evaluated, and hence there was expectancy to detect a smell with presentation of 0.06% n-butanol, a concentration much more difficult to detect than the 0.32% concentration used at the second preoperative site. Subsequent presentation of the highest concentration (3.2%) resulted in differential detection rates that were associated with the concentration of the first stimulus, i.e., apparently if a subject had to “work harder” to detect the first odor, subjects more readily detected the subsequent high concentration stimulus and hence were less likely to be characterized as anosmic. Despite this apparent difference in sensitivity attributable to concentration test sequence, a putative false positive anosmia identification rate at the second site is in fact a categorical demonstration of “severe hyposmia or anosmia”, i.e., a failure to detect 3.2% n-butanol is unambiguously a severe impairment. Therefore, we suggest that any patient, regardless of age, that presents with a sudden onset of dysosmia, which cannot be otherwise explained, should be immediately tested for COVID-19 infection.

Two-odorant concentration tests of olfactory sensitivity reliably detects both modest and severe olfactory impairments as a function of age and sex, and could be used in the context of supply shortages during a global pandemic. Such odorant sensitivity tests are not diagnostics, but a quick screening method that can be used to identify patients highly likely to be infected and that also provides the benefits of result immediacy and high frequency repeated testing at very low cost (38, 39). Widespread frequent screening for olfactory impairment in the general population may be used as an indicator of COVID-19 spread, is sensitive to lockdown interventions, and might assist public health monitoring and interventions (40). Manufacturing and mass deployment of crushable ampules or other packaging systems are possible and could greatly increase our understanding of prevalence and risk factors once reporting systems are in place.

With variants that present patients with a high likelihood of olfactory impairment, use in screening and health care contexts could identify those COVID-19 patients at greater risk of severe disease and prompt consideration of immediate medical interventions. If used broadly in conjunction with rapid screening (antigen) tests, viral positivity in conjunction with normal olfaction would enable more rapid recognition and medical intervention for those patients at risk of severe disease and a poorer prognosis.

## Data Availability

All data produced in the present study are available upon reasonable request to the authors

## Acknowledgements

Initial use was undertaken by the staff of the UR Primary Care Network Central Respiratory Clinic, Wallace Johnson MD director; subsequent testing was performed by the UR Medicine Ambulatory Pre-Procedure Testing staff at the River Road and Sawgrass sites. Initial consultation with Ann M. Dozier PhD and Shawn Newlands, M.D., Ph.D., M.B.A. appreciated. Crushable ampules provided by James Alexander Corporation, Blairstown NJ. US Patent Application US 2022/0054074 A1 was published 24 February 2022.

## Supporting information

**S1 Fig. URSmellTest™**. Two ampules of 0.32 and 3.2% n-butanol in water conjoined with clear polyolefin shrink tubing that overlaps the central glass crush area. Top: before use; Bottom: crushed.

**S2 Fig. Instruction sheet used in the clinical setting to standardize testing procedures**

**S1 Table. Ethnic and racial demographics of 2450 patients presenting to the preprocedural testing clinics that were approached for olfactory testing with n-butanol in crushable ampules**. No patients were excluded from this tabulation that extended several days after the retrieval of the data used for logistic regression.

**S2 Table. Estimated incidence of dysosmia as a function of subject age and odorant concentration**. Linear regression was performed on the coefficients and constants of three logistic regressions in table 1 (0.06%, 0.32%, and 3.2% from site 2) as a function of the log of test concentration. These two linear regressions1 were used to obtain coefficients and constants for intermediate concentrations and were then used to calculate prevalence estimates as a function of age. Hyposmia occurs in the tail of the distribution of olfactory sensitivity in the population distribution and is a joint function of age and odorant concentration; thus, if hyposmia is defined as the least sensitive 12% of a sample, test concentrations can be selected for different geriatric or education settings. For example, 1% might be selected for a population over the age of 40 and would be expected to approximate the NHANEs (1) hyposmia data (Fig 1) generated using odor identification error rates; 0.56% might be used in college and university settings.

## Notes

### Competing Interest Statement

The authors have declared no competing interest.

### Funding Statement

This study did not receive any funding

### Summary of Updates

Independent logistic regression analyses were undertaken for both sexes at 0.32 and 3.2% n-butanol. A sex difference at advanced age was observed at the higher concentration and is reported in the figure and associated table. Title and abstract changed accordingly; new findings reported and discussed.

## References

1. Hoffman HJ, Rawal S, Li C-M, Duffy VB. New chemosensory component in the U.S. National Health and Nutrition Examination Survey (NHANES): first-year results for measured olfactory dysfunction. Rev Endocr Metab Disord. 2016;17(2):221–40.

2. Agyeman AA, Chin KL, Landersdorfer CB, Liew D, Ofori-Asenso R. Smell and Taste Dysfunction in Patients With COVID-19: A Systematic Review and Meta-analysis. Mayo Clin Proc. 2020;95(8):1621–31.

3. Kim DH, Kim MG, Yang SJ, Lee EJ, Yeom SW, You YS, et al. Influenza and anosmia: Important prediction factors for severity and death of COVID-19. J Infect. 2021.

4. Borsetto D, Hopkins C, Philips V, Obholzer R, Tirelli G, Polesel J, et al. Self-reported alteration of sense of smell or taste in patients with COVID-19: a systematic review and meta-analysis on 3563 patients. Rhinology. 2020;58(5):430–6.

5. Houser D. Less is More: COVID-19 Illness Severity Negatively Related to Smell and Taste Loss. Utrecht: Utrecht University; 2020.

6. Izquierdo-Dominguez A, Rojas-Lechuga MJ, Chiesa-Estomba C, Calvo-Henriquez C, Ninchritz-Becerra E, Soriano-Reixach M, et al. Smell and Taste Dysfunction in COVID-19 Is Associated With Younger Age in Ambulatory Settings: A Multicenter Cross-Sectional Study. J Investig Allergol Clin Immunol. 2020;30(5):346–57.

7. Coelho DH, Reiter ER, Budd SG, Shin Y, Kons ZA, Costanzo RM. Predictors of smell recovery in a nationwide prospective cohort of patients with COVID-19. Am J Otolaryngol. 2021;43(1):103239.

8. Yan CH, Faraji F, Prajapati DP, Ostrander BT, DeConde AS. Self-reported olfactory loss associates with outpatient clinical course in COVID-19. Int Forum Allergy Rhinol. 2020;10(7):821–31.

9. Vaira LA, Lechien JR, Khalife M, Petrocelli M, Hans S, Distinguin L, et al. Psychophysical Evaluation of the Olfactory Function: European Multicenter Study on 774 COVID-19 Patients. Pathogens. 2021;10(1).

10. Sayin P, Altinay M, Cinar AS, Ozdemir HM. Taste and Smell Impairment in Critically Ill Patients With COVID-19: An Intensive Care Unit Study. Ear Nose Throat J. 2020:145561320977464.

11. Barry M, AlMohaya A, AlHijji A, Akkielah L, AlRajhi A, Almajid F, et al. Clinical Characteristics and Outcome of Hospitalized COVID-19 Patients in a MERS-CoV Endemic Area. J Epidemiol Glob Health. 2020;10(3):214–21.

12. Kaeuffer C, Le Hyaric C, Fabacher T, Mootien J, Dervieux B, Ruch Y, et al. Clinical characteristics and risk factors associated with severe COVID-19: prospective analysis of 1,045 hospitalised cases in North-Eastern France, March 2020. Euro Surveill. 2020;25(48).

13. Lechien JR, Chiesa-Estomba CM, Varia LA, De Riu G, Cammaroto G, Chekkoury-Idrissi Y, et al. Epidemiological, otolaryngological, olfactory and gustatory outcomes according to the severity of COVID-19: a study of 2579 patients. Eur Arch Otorhinolaryngol. 2021.

14. Purja S, Shin H, Lee JY, Kim E. Is loss of smell an early predictor of COVID-19 severity: a systematic review and meta-analysis. Arch Pharm Res. 2021.

15. Talavera B, Garcia-Azorin D, Martinez-Pias E, Trigo J, Hernandez-Perez I, Valle-Penacoba G, et al. Anosmia is associated with lower in-hospital mortality in COVID-19. J Neurol Sci. 2020;419:117163.

16. Hopkins C, Vaira LA, De Riu G. Self-reported olfactory loss in COVID-19: is it really a favorable prognostic factor? Int Forum Allergy Rhinol. 2020;10(7):926.

17. Vaira LA, Salzano G, De Riu G. The importance of olfactory and gustatory disorders as early symptoms of coronavirus disease (COVID-19). Br J Oral Maxillofac Surg. 2020;58(5):615–6.

18. Vaira LA, Hopkins C, Petrocelli M, Lechien JR, Soma D, Giovanditto F, et al. Do olfactory and gustatory psychophysical scores have prognostic value in COVID-19 patients? A prospective study of 106 patients. J Otolaryngol Head Neck Surg. 2020;49(1):56.

19. Eliezer M, Hamel AL, Houdart E, Herman P, Housset J, Jourdaine C, et al. Loss of smell in patients with COVID-19: MRI data reveal a transient edema of the olfactory clefts. Neurology. 2020;95(23):e3145–e52.

20. Hornuss D, Lange B, Schroter N, Rieg S, Kern WV, Wagner D. Anosmia in COVID-19 patients. Clin Microbiol Infect. 2020.

21. Pang KW, Chee J, Subramaniam S, Ng CL. Frequency and Clinical Utility of Olfactory Dysfunction in COVID-19: a Systematic Review and Meta-analysis. Curr Allergy Asthma Rep. 2020;20(12):76.

22. Hannum ME, Ramirez VA, Lipson SJ, Herriman RD, Toskala AK, Lin C, et al. Objective Sensory Testing Methods Reveal a Higher Prevalence of Olfactory Loss in COVID-19-Positive Patients Compared to Subjective Methods: A Systematic Review and Meta-Analysis. Chem Senses. 2020;45(9):865–74.

23. Kobayashi M, Saito S, Kobayakawa T, Deguchi Y, Costanzo RM. Cross-cultural comparison of data using the odor stick identification test for Japanese (OSIT-J). Chem Senses. 2006;31(4):335–42.

24. Moein ST, Hashemian SM, Tabarsi P, Doty RL. Prevalence and reversibility of smell dysfunction measured psychophysically in a cohort of COVID-19 patients. Int Forum Allergy Rhinol. 2020;10(10):1127–35.

25. Niklassen AS, Ovesen T, Fernandes H, Fjaeldstad AW. Danish validation of sniffin’ sticks olfactory test for threshold, discrimination, and identification. Laryngoscope. 2018;128(8):1759–66.

26. Konstantinidis I, Printza A, Genetzaki S, Mamali K, Kekes G, Constantinidis J. Cultural adaptation of an olfactory identification test: the Greek version of Sniffin’ Sticks. Rhinology. 2008;46(4):292–6.

27. Tekeli H, Altundag A, Salihoglu M, Cayonu M, Kendirli MT. The applicability of the “Sniffin’ Sticks” olfactory test in a Turkish population. Med Sci Monit. 2013;19:1221–6.

28. Ribeiro JC, Simoes J, Silva F, Silva ED, Hummel C, Hummel T, et al. Cultural Adaptation of the Portuguese Version of the “Sniffin’ Sticks” Smell Test: Reliability, Validity, and Normative Data. PLoS One. 2016;11(2):e0148937.

29. Oleszkiewicz A, Taut M, Sorokowska A, Radwan A, Kamel R, Hummel T. Development of the Arabic version of the “Sniffin’ Sticks” odor identification test. Eur Arch Otorhinolaryngol. 2016;273(5):1179–84.

30. Doty RL, Shaman P, Dann M. Development of the University of Pennsylvania Smell Identification Test: a standardized microencapsulated test of olfactory function. Physiol Behav. 1984;32(3):489–502.

31. Hummel T, Sekinger B, Wolf SR, Pauli E, Kobal G. ‘Sniffin’ sticks’: olfactory performance assessed by the combined testing of odor identification, odor discrimination and olfactory threshold. Chem Senses. 1997;22(1):39–52.

32. Marchese-Ragona R, Restivo DA, De Corso E, Vianello A, Nicolai P, Ottaviano G. Loss of smell in COVID-19 patients: a critical review with emphasis on the use of olfactory tests. Acta Otorhinolaryngol Ital. 2020;40(4):241–7.

33. Zernecke R, Vollmer B, Albrecht J, Kleemann AM, Haegler K, Linn J, et al. Comparison of two different odorants in an olfactory detection threshold test of the Sniffin’ Sticks. Rhinology. 2010;48(3):368–73.

34. Kingdom S. Logistic Regression Calculator [Available from: https://www.statskingdom.com/420logistic_regression.html.

35. Blue S. Logistic Regression Calculator [Available from: https://stats.blue/Stats_Suite/logistic_regression_calculator.html.

36. Kobal G, Klimek L, Wolfensberger M, Gudziol H, Temmel A, Owen CM, et al. Multicenter investigation of 1,036 subjects using a standardized method for the assessment of olfactory function combining tests of odor identification, odor discrimination, and olfactory thresholds. Eur Arch Otorhinolaryngol. 2000;257(4):205–11.

37. Cometto-Muniz JE, Abraham MH. Human olfactory detection of homologous n-alcohols measured via concentration-response functions. Pharmacol Biochem Behav. 2008;89(3):279–91.

38. Larremore DB, Wilder B, Lester E, Shehata S, Burke JM, Hay JA, et al. Test sensitivity is secondary to frequency and turnaround time for COVID-19 surveillance. medRxiv. 2020.

39. Mina MJ, Parker R, Larremore DB. Rethinking Covid-19 Test Sensitivity - A Strategy for Containment. N Engl J Med. 2020.

40. Pierron D, Pereda-Loth V, Mantel M, Moranges M, Bignon E, Alva O, et al. Smell and taste changes are early indicators of the COVID-19 pandemic and political decision effectiveness. Nat Commun. 2020;11(1):5152.

