## Supplementary figures and images for "Detection of COVID-19 and age-dependent dysosmia with paired crushable odorant ampules"

### Supplemental Figure 1

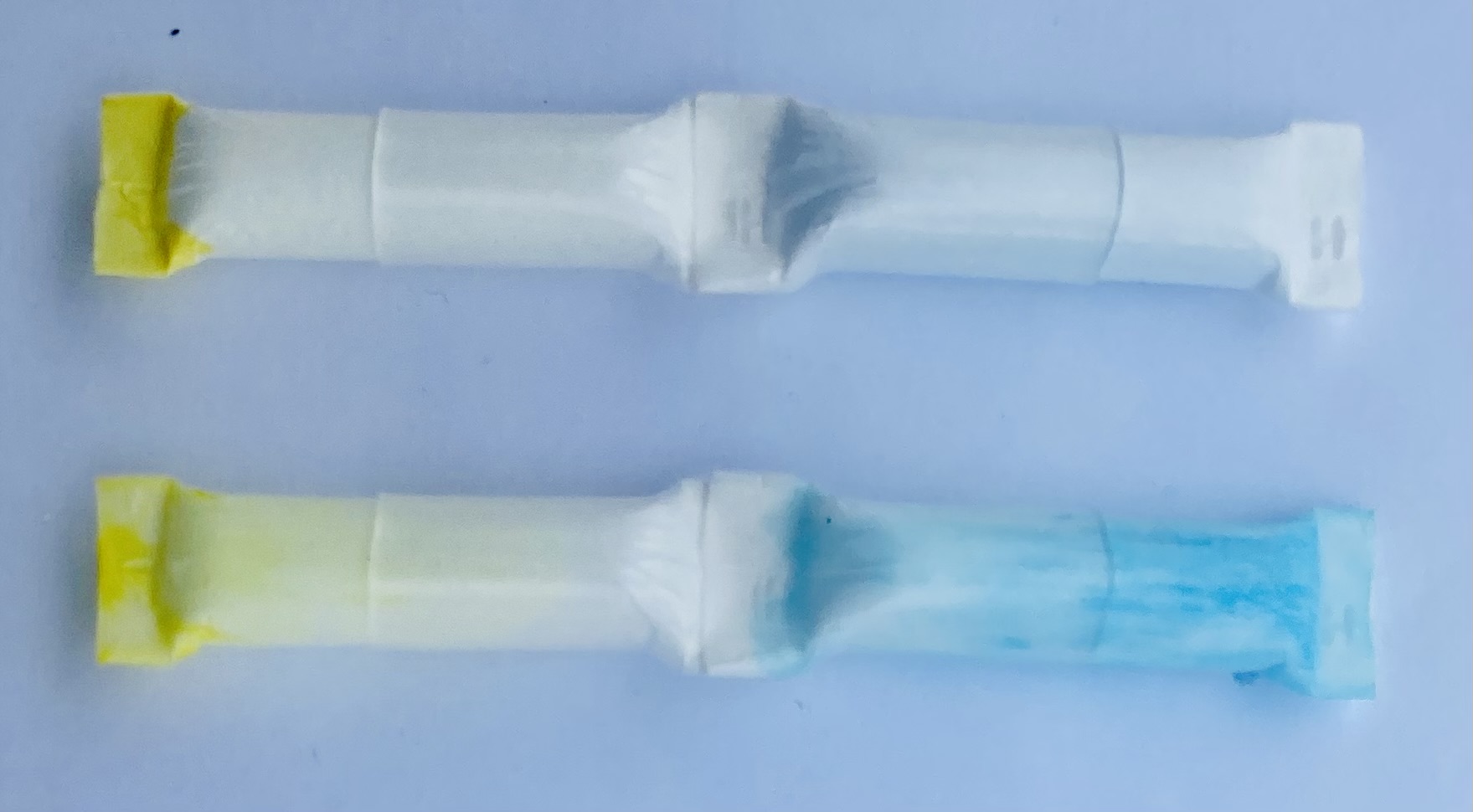
