## Supplemental Figure 2 for "Detection of COVID-19 and age-dependent dysosmia with paired crushable odorant ampules"

### How to use *yoUR* Smell Test

#### TEST ADMINISTRATION

1) Select a UR smell test kit, hold it between your thumb and forefinger, with your arm at your side.
 Say: "**We're going to test your ability to smell now**"

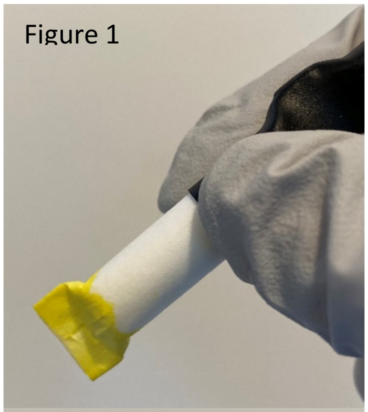
Figure 1

2) Bring the uncrushed ampule up to the patient's nose and ask:

**"While inhaling deeply through your nose, do you smell anything?"**

While demonstrating strong sniffing for the patient (as if you have a runny nose.) **At this point, the patient may indicate they smell the gloves, perfumed soap/sanitizers, ambient smells in the room.*

3) Lower the ampule with yellow end ("L" embossed on end) facing down **(Figure 1)** crush the ampule ONCE toward the middle, it will make a popping noise and the white case will turn yellow after a few seconds as you keep the yellow end oriented downwards. **(Figure 2)**.

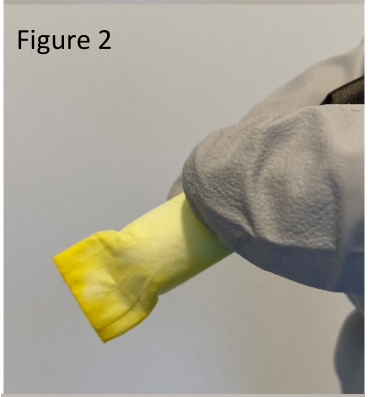
Figure 2

4) **After 4 to 5 seconds**, bring the crushed ampule to the patient's nose, and ask:
**"Do you smell anything?"**
Note the response.
(Yes: likely normal smell; No: hyposmia or anosmia depends on next challenge)

5) Lower the ampule and rotate the test with the opposite end down ("H" embossed on end) **(Figure 3).** Crush the ampule ONCE towards the middle, it will make a popping noise and the white case will **turn blue** after a few seconds.

**After 4 to 5 seconds**, bring the crushed ampule to the patient's nose and ask:
**"Do you smell anything?"**
Note the response.
(Yes: Normal or Hyposmic [cannot smell low concentration]; No: Anosmic)

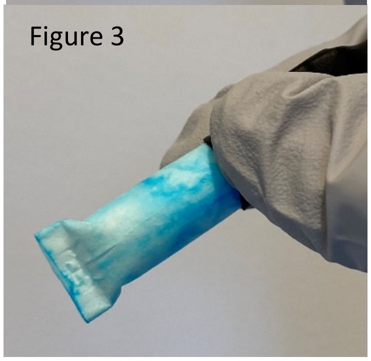
Figure 3

#### INTERPRETATION OF RESULTS

A person with a normal sense of smell should respond:
- No to blank (If they note the ambient smells in the room that is OK)
- Yes to low concentration (e.g., "Different, but something is there.", "Hard to smell, but I do smell something."…)
- Yes to high concentration (e.g., "Definitely different", "More intense" may also have a strong response with facial expression, turning head away.)

An anosmic person can't smell anything.
An hyposmic person cannot smell the low concentration (yellow) but can smell the high concentration (blue).

This test assesses sensitivity to n-butanol, a non-toxic food additive in water with added food coloring.

Present Uncrushed Ampule

**While inhaling deeply through your nose, do you smell anything?**

**No**

Crush, wait 4-5 seconds
Present crushed Low Concentration

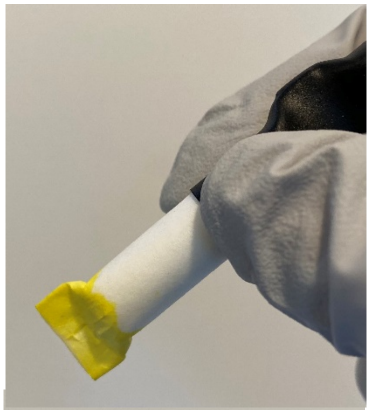

If patient smells the blank, Discuss gloves, plastic, perfume or other items in room; repeat

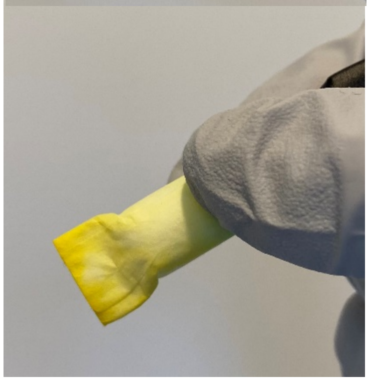

**Do you smell anything?**

**Yes: Normal**

Present crushed High Concentration

**No: anosmic or hyposmic**

Crush, wait 4-5 seconds
Present crushed High Concentration

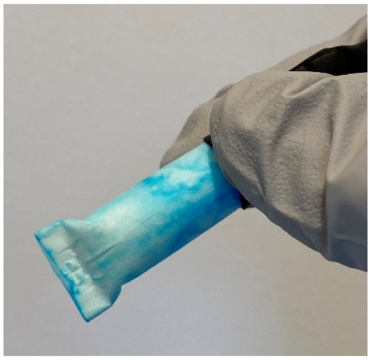

**Do you smell anything?**

**No: Anosmic**

**Yes: Hyposmic**

**Do you smell anything?**

**Yes: Normal**

**No: Error, Repeat test**

This test assesses sensitivity to n-butanol, a non-toxic food additive in water with added food coloring.
