## Supplemental Table 1 for "Detection of COVID-19 and age-dependent dysosmia with paired crushable odorant ampules"

**S1 Table. Ethnic and racial demographics of 2450 patients presenting to the preprocedural testing clinics that were approached for olfactory testing with n-butanol in crushable ampules.** No patients were excluded from this tabulation that extended several days after the retrieval of the data used for logistic regression.

| Ethnicity | Site 1 | Site 2 | Total |
|---|---|---|---|
| Hispanic or Latinx | 54 | 94 | 148 |
| Not Hispanic or Latinx | 821 | 1247 | 2068 |
| Unknown | 74 | 160 | 234 |
| Total | 949 | 1501 | 2450 |

| Race | | | |
|---|---|---|---|
| American Indian or Alaska Native | 2 | 3 | 5 |
| Asian | 11 | 23 | 34 |
| Black or African American | 113 | 226 | 339 |
| Native Hawaiian or Other Pacific Islander | 1 | 0 | 1 |
| White or Caucasian | 770 | 1142 | 1912 |
| Multiracial/Other | 39 | 72 | 111 |
| Unknown | 13 | 35 | 48 |
| Total | 949 | 1501 | 2450 |
