## Supplemental Table 2 for "Detection of COVID-19 and age-dependent dysosmia with paired crushable odorant ampules"

**S2 Table. Estimated incidence of dysosmia as a function of subject age and odorant concentration.** Linear regression was performed on the coefficients and constants of three logistic regressions in table 1 (0.06%, 0.32%, and 3.2% from site 2) as a function of the log of test concentration. These two linear regressions[1] were used to obtain coefficients and constants for intermediate concentrations and were then used to calculate prevalence estimates as a function of age. Hyposmia occurs in the tail of the distribution of olfactory sensitivity in the population distribution and is a joint function of age and odorant concentration; thus, if hyposmia is defined as the least sensitive 12% of a sample, test concentrations can be selected for different geriatric or education settings. For example, 1% might be selected for a population over the age of 40 and would be expected to approximate the NHANEs (1) hyposmia data (Fig 1) generated using odor identification error rates; 0.56% might be used in college and university settings.

| Age | Odorant Concentration (% n-butanol in water) | | | | | | | |
|---|---|---|---|---|---|---|---|---|
| | 3.2 | 1.8 | 1.0 | 0.56 | 0.32 | 0.18 | 0.1 | 0.056 |
| **5** | 0.006 | 0.025 | 0.056 | 0.085 | 0.107 | **0.122** | 0.131 | 0.137 |
| 10 | 0.007 | 0.029 | 0.062 | 0.093 | 0.116 | 0.131 | 0.14 | 0.146 |
| **15** | 0.009 | 0.033 | 0.069 | 0.101 | **0.125** | 0.14 | 0.15 | 0.155 |
| 20 | 0.011 | 0.038 | 0.076 | 0.11 | 0.134 | 0.15 | 0.16 | 0.165 |
| **25** | 0.013 | 0.043 | 0.084 | **0.12** | 0.144 | 0.161 | 0.17 | 0.176 |
| 30 | 0.016 | 0.05 | 0.093 | 0.13 | 0.155 | 0.172 | 0.182 | 0.187 |
| 35 | 0.019 | 0.057 | 0.103 | 0.141 | 0.167 | 0.183 | 0.193 | 0.199 |
| 40 | 0.023 | 0.065 | 0.114 | 0.153 | 0.179 | 0.195 | 0.205 | 0.211 |
| **45** | 0.028 | 0.074 | **0.126** | 0.166 | 0.192 | 0.208 | 0.218 | 0.224 |
| 50 | 0.034 | 0.085 | 0.139 | 0.18 | 0.205 | 0.222 | 0.232 | 0.237 |
| 55 | 0.041 | 0.097 | 0.153 | 0.194 | 0.22 | 0.236 | 0.245 | 0.251 |
| 60 | 0.05 | 0.11 | 0.168 | 0.209 | 0.235 | 0.251 | 0.26 | 0.265 |
| **65** | 0.06 | **0.125** | 0.184 | 0.225 | 0.25 | 0.266 | 0.275 | 0.28 |
| 70 | 0.072 | 0.141 | 0.202 | 0.242 | 0.267 | 0.282 | 0.29 | 0.295 |
| 75 | 0.086 | 0.159 | 0.22 | 0.26 | 0.284 | 0.298 | 0.306 | 0.311 |
| 80 | 0.103 | 0.18 | 0.24 | 0.279 | 0.301 | 0.315 | 0.323 | 0.327 |
| **85** | **0.123** | 0.202 | 0.261 | 0.298 | 0.32 | 0.332 | 0.34 | 0.344 |
| 90 | 0.146 | 0.226 | 0.283 | 0.318 | 0.338 | 0.35 | 0.357 | 0.361 |
| 95 | 0.173 | 0.252 | 0.306 | 0.339 | 0.358 | 0.369 | 0.375 | 0.379 |
| 100 | 0.203 | 0.28 | 0.331 | 0.361 | 0.377 | 0.387 | 0.393 | 0.396 |

[1] $\beta_0$: slope -1.086, intercept -1.86
$\beta_1$: slope .00785, intercept 0.145
